# Lessons learned and lessons missed: Impact of the Covid-19 pandemic on all-cause mortality in 40 industrialised countries prior to mass vaccination

**DOI:** 10.1101/2021.07.12.21260387

**Authors:** Vasilis Kontis, James E Bennett, Robbie M Parks, Theo Rashid, Jonathan Pearson-Stuttard, Perviz Asaria, Bin Zhou, Michel Guillot, Colin D Mathers, Young-Ho Khang, Martin McKee, Majid Ezzati

## Abstract

Industrialised countries have varied in their early response to the Covid-19 pandemic, and how they have adapted to new situations and knowledge since the pandemic began. These variations in preparedness and policy may lead to different death tolls from Covid-19 as well as from other diseases. We applied an ensemble of 16 Bayesian probabilistic models to vital statistics data to estimate the impacts of the pandemic on weekly all-cause mortality for 40 industrialised countries from mid-February 2020 through mid-February 2021, before a large segment of the population was vaccinated in any of these countries. Taken over the entire year, an estimated 1,401,900 (95% credible interval 1,259,700-1,572,500) more people died in these 40 countries than would have been expected had the pandemic not taken place. This is equivalent to 140 (126-157) additional deaths per 100,000 people and a 15% (13-17) increase in deaths over this period in all of these countries combined. In Iceland, Australia and New Zealand, mortality was lower over this period than what would be expected if the pandemic had not occurred, while South Korea and Norway experienced no detectable change in mortality. In contrast, the populations of the USA, Czechia, Slovakia and Poland experienced at least 20% higher mortality. There was substantial heterogeneity across countries in the dynamics of excess mortality. The first wave of the pandemic, from mid-February to the end of May 2020, accounted for over half of excess deaths in Scotland, Spain, England and Wales, Canada, Sweden, Belgium and Netherlands. At the other extreme, the period between mid-September 2020 and mid-February 2021 accounted for over 90% of excess deaths in Bulgaria, Croatia, Czechia, Hungary, Latvia, Montenegro, Poland, Slovakia and Slovenia. Until the great majority of national and global populations have vaccine-acquired immunity, minimising the death toll of the pandemic from Covid-19 and other diseases will remain dependent on actions to delay and contain infections and continue routine health and social care.

---

Many industrialised countries experienced a rise in all-cause mortality in the first wave of the Covid-19 pandemic, while others avoided any excess deaths^1^. These excess deaths were due to infection with SARS-CoV-2, delays and disruptions in the provision and use of healthcare for other diseases, loss of jobs and income, disruptions of social networks and support, and changes in nutrition, drug and alcohol use, transportation, crime, and violence^2, 3^.

Decline in infections following initial lockdowns and other restrictions, and advances in knowledge about the SARS-CoV-2 transmission and infection, presented a window of opportunity for countries to implement pandemic control measures and strengthen health and social care provision that would minimise the impacts of subsequent waves^4, 5^. Comparative analysis of excess deaths helps understand how effectively these measures were implemented and how resilient the health and social care system was in each country. We quantified the weekly mortality impacts of the first year of the Covid-19 pandemic, from mid-February 2020 to mid-February 2021, in 40 industrialised countries, listed below. We used this period because mortality due to the pandemic was negligible before mid-February 2020^1^, and vaccination rates against SARS-CoV-2 were still relatively low before mid-February 2021 in these countries (no more than 4% of the population had received both doses in any of these countries). After mid-February 2021, the effect of vaccines on mortality was expected to appear in some countries, which should be subject to a distinct analysis.

We selected countries for our analysis if their total population in 2020 was more than 100,000 and if we could access weekly data on all-cause mortality that went back at least to 2016 and extended through mid-February 2021. The 40 countries in our analysis were divided into five geographical regions: the Pacific (Australia, New Zealand, South Korea), the Americas (Canada, Chile, the USA), Central and Eastern Europe (Bulgaria, Croatia, Czechia, Estonia, Hungary, Latvia, Lithuania, Montenegro, Poland, Romania, Serbia, Slovakia, Slovenia), Southwestern Europe (Cyprus, France, Greece, Italy, Malta, Portugal, Spain), Northwestern Europe (Austria, Belgium, England and Wales, Germany, Luxembourg, the Netherlands, Northern Ireland, Scotland, Switzerland) and Nordic (Denmark, Finland, Iceland, Norway, Sweden). In addition to national estimates, we separately estimates excess deaths for all 50 US states and the District of Columbia, some of which are larger than most other countries included in our analysis, because the extent and temporal dynamics of the pandemic were heterogeneous across states.

We used a probabilistic model averaging approach, using an ensemble of 16 Bayesian models, to estimate what death rates were expected to be over this period had the pandemic not occurred, and compared these estimates with actual deaths from all causes in each country. The analytical method was designed to enhance comparison across countries and over time, and account for medium-long-term secular trends in mortality, the potential dependency of death rates in each week on those in preceding week(s) and in each year on those in preceding year(s), and factors that affect mortality including seasonality, temperature and public holidays.

We used data on weekly deaths from the start of time series of data through mid-February 2020 to estimate the parameters of each model, which were then used to predict death rates for the subsequent 52 weeks as estimates of how many deaths would have occurred without the pandemic. These were then compared to reported deaths to calculate excess mortality due to the pandemic. We report the number of excess deaths, excess deaths per 100,000 people, and relative (percent) increase in deaths together with their corresponding 95% credible intervals. For the purpose of reporting, we rounded results on number of deaths that are 1,000 or more to the nearest hundred to avoid giving a false sense of precision in the presence of uncertainty; results less than 1,000 were rounded to the nearest ten. We also report the posterior probability that an estimated change in deaths corresponds to a true increase (or decrease), as described in Methods. We report results for the entire year, as well as for three non-overlapping periods: the first wave of the pandemic (from mid-February 2020 through end of May), the (northern hemisphere) summer period (from beginning of June to mid-September 2020) and subsequent wave(s) (from mid-September 2020, when schools normally open in the northern hemisphere, to mid-February 2021).

Taken over the entire year, both sexes and all ages, an estimated 1,401,900 (95% credible interval 1,259,700-1,572,500) more people died in these 40 countries than would have been expected had the pandemic not taken place. This is equivalent to 140 (126-157) additional deaths per 100,000 people and a 15% (13-17) increase in deaths over this period in all of these countries combined. The number of deaths assigned to Covid-19 in these countries over the same period was 1,253,846, which is 90% of the excess all-cause death toll (Extended Data Table 1). The number of excess deaths were largest in the USA (621,100; 520,100-749,700), followed by Italy (118,100; 87,300-148,000) and England and Wales (102,100; 75,300-129,000) (Fig. 1 and Extended Data Table 1). Within the USA, California (71,900; 64,100-79,600) and Texas (57,600; 48,300-67,500) experienced the largest number of excess deaths, about the same as excess deaths in Spain and France, respectively (Extended Data Fig. 3).

**Fig. 1.**
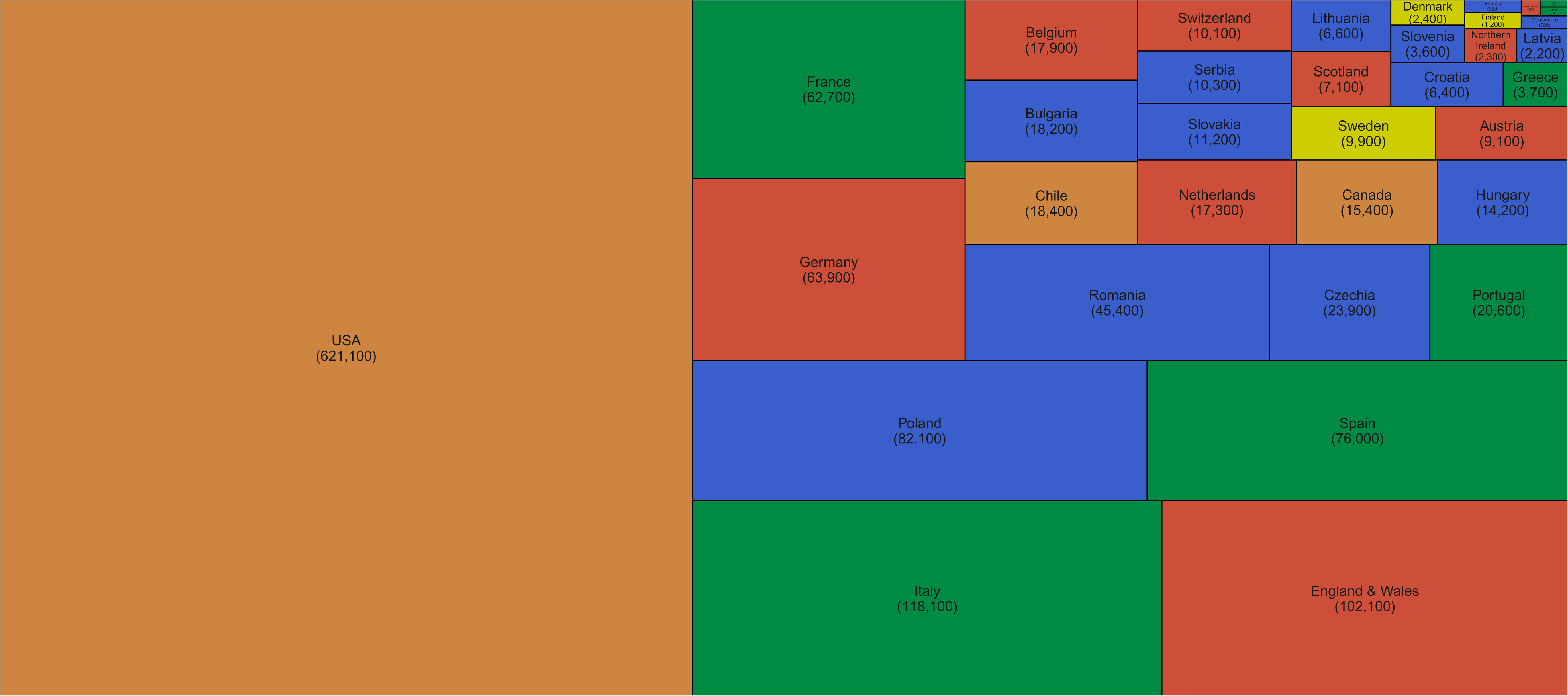
Number of excess deaths due to the first year of the Covid-19 pandemic by country. The size of each rectangle shows the number of deaths from all causes in excess of what would be expected if there had been no Covid-19 pandemic from mid-February 2020 through mid-February 2021 for each country. There are no segments for Australia, New Zealand, Norway, Iceland and South Korea because we estimated no detectable excess deaths or a potential reduction in mortality compared to the no-pandemic baseline. Colour for each country indicates its geographical region as stated in the Text. See Extended Data Fig. 3 for results for US states.

In Iceland, Australia and New Zealand, mortality was 3-6% lower over this period than what would be expected if the pandemic had not occurred, with posterior probabilities of the estimated decrease being a true decrease ranging 92-95% (Fig. 2). South Korea and Norway experienced no detectable change in mortality (53% and 74% probability of an increase respectively, with posterior median estimated increases <2%), and Finland, Greece, Cyprus and Denmark experienced increases of 2-5% (Fig. 2A), with posterior probabilities that these changes represent an increase in death ranging from 80% to 97%. At the other extreme, the populations of the USA, Czechia, Slovakia and Poland experienced at least 20% higher mortality over these 52 weeks than they would have had the pandemic not occurred; the increase was between 15% and 20% in England and Wales, Spain, Italy, Portugal, Romania, Lithuania, Bulgaria, Slovenia, Chile, Belgium and Switzerland; the posterior probabilities that these countries experienced an increase in deaths were >99%. Because baseline mortality (i.e., death rates expected without the pandemic) varied across countries, the ordering of countries in terms of excess deaths per 100,000 people (Fig. 2B) differed from the ranking of percent increase. Bulgaria, Lithuania, Romania, Czechia, Poland, Slovakia and Portugal experienced more than 200 excess deaths per 100,000 people and Italy, USA, Spain, Slovenia, England and Wales, Belgium and Croatia between 150 and 200, all with posterior probabilities of an increase in deaths >99%. There was as much variation in excess mortality across US states as across the 40 countries together, with Hawaii having experienced the same level of mortality as would have been expected without the pandemic, Maine a 5% increase, and, at the other extreme, New Jersey, Arizona, Mississippi, Texas, California, Louisiana and New York at least 25% higher mortality over this year (Extended Data Fig. 4).

**Fig. 2.**
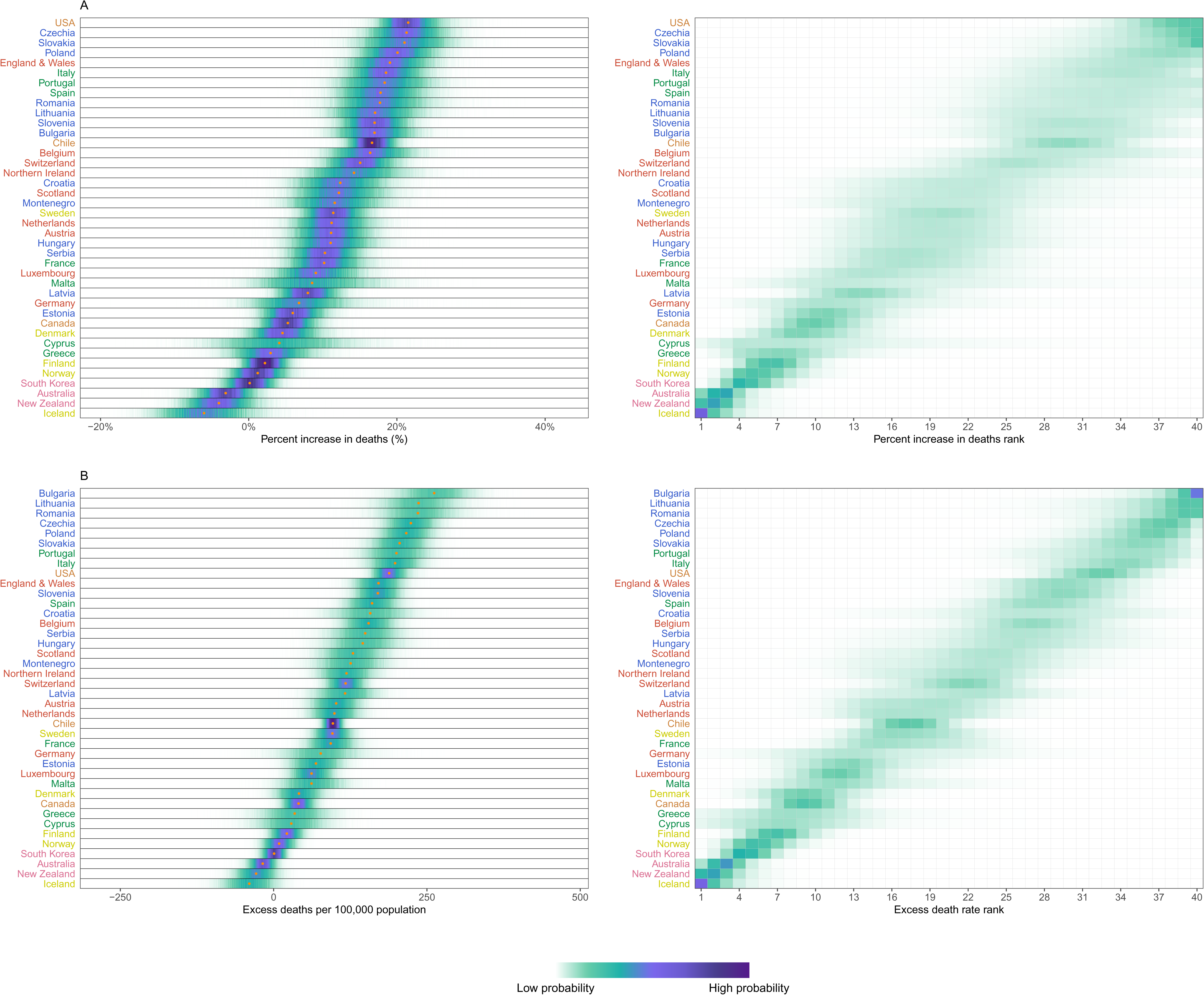
Excess mortality due to the Covid-19 pandemic, by country. (A) Posterior distribution of percent increase in deaths from any cause from mid-February 2020 to mid-February 2021. Gold dots show the posterior medians. (B) Posterior distribution of excess deaths from any cause per 100,000 people from mid-February 2020 to mid-February 2021. Gold dots show the posterior medians. In both panels, the right hand side shows the probability distribution for the country’s rank. Countries are ordered vertically by median increase from smallest (at the bottom) to the largest (at the top). Colour for each country’s name indicates its geographical region as stated in the Text. See Extended Data Fig. 6 for results by sex. See Extended Data Fig. 4 for results for US states.

There was substantial heterogeneity across countries in terms of the patterns and dynamics of excess mortality over time (Extended Data Figures 1 and 2). Some countries in Central and Eastern Europe – Bulgaria, Lithuania, Poland, Romania, Serbia and Montenegro – had no or little excess mortality in the first wave of the pandemic (mid-February 2020 to end of May 2020), but experienced between 5% and 13% increase in mortality during the (northern hemisphere) summer (June 2020 to mid-September 2020; Fig. 3A). In contrast, some countries with medium to high levels of excess mortality in the first wave returned to death rates in the summer that were about the same as the no-pandemic baseline (England and Wales, Belgium, Scotland, Northern Ireland, Sweden, Netherlands, France, Canada, Switzerland, Luxembourg and Cyprus) or only slightly higher than this baseline (Italy and Spain). Portugal and the USA experienced a similar increase in mortality over the summer – 10% (1-21) and 17% (12-24), respectively – to what they had in the first wave. During the same period, Australia, New Zealand and Iceland had a mortality deficit compared to levels that would have been expected without a pandemic. In Australia and New Zealand, which were in winter season in this period, this reduction has been attributed to fewer deaths from seasonal flu due to reduced contact among people ^6–9^. Chile, the other southern hemisphere country in our analysis, had 12% (8-17) higher mortality in the first wave, followed by an even larger increase of 21% (15-26) during the (southern hemisphere) winter period.

**Fig. 3.**
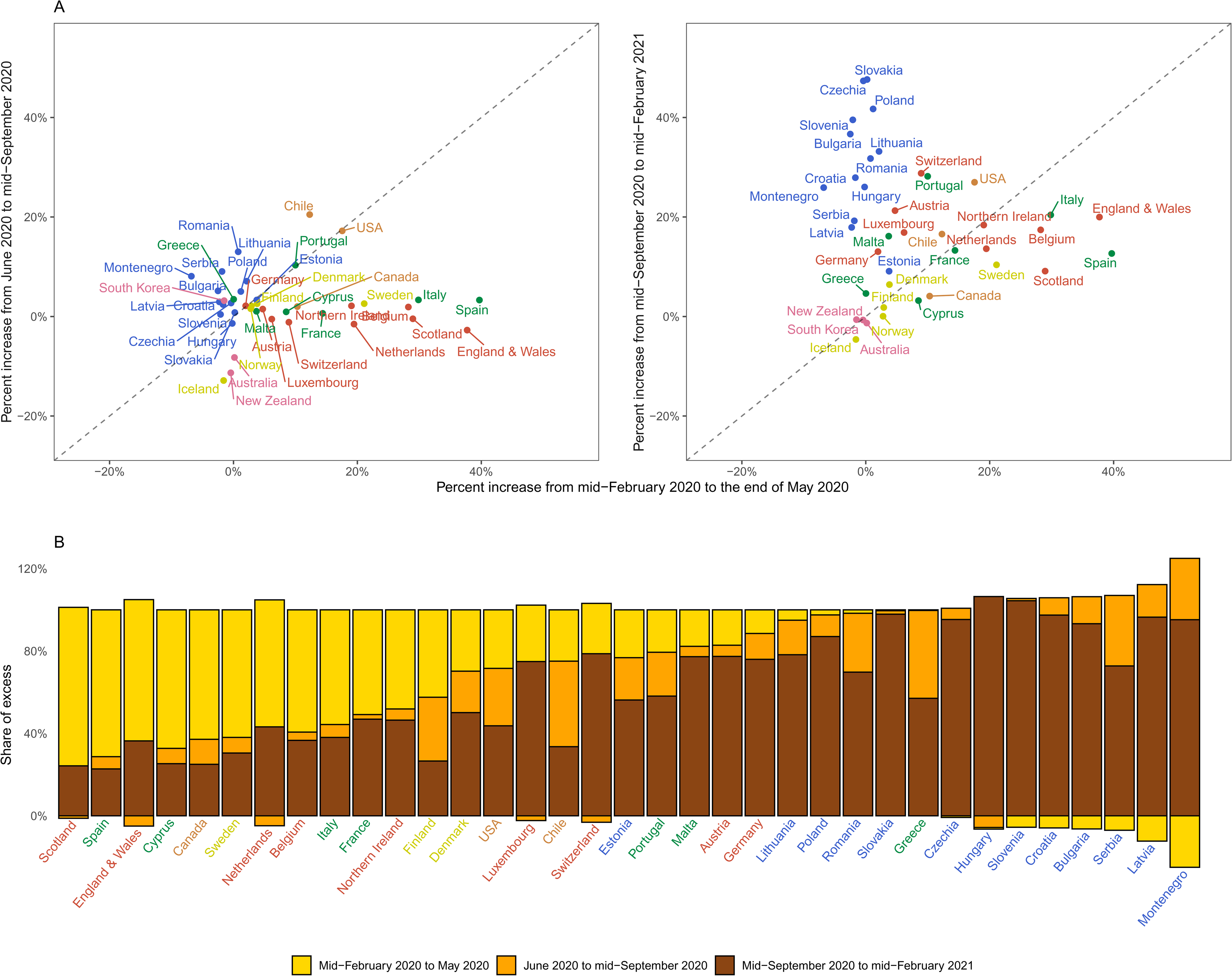
Excess mortality due to the Covid-19 pandemic in different time periods. (A) Comparison of percent increase in mortality from any cause in excess of what would be expected if there had been no Covid-19 pandemic in summer (beginning of June 2020 to mid-September 2020) and subsequent waves (mid-September 2020 to mid-February 2021) with the first wave (mid-February 2020 to end of May 2020) in each country. (B) Proportion of excess deaths in each of the above three periods in each country. There are no bars for Australia, New Zealand, Norway, Iceland and South Korea in panel B because we estimated no detectable excess deaths or a potential reduction in mortality compared to the no-pandemic baseline. Colour for each country indicates its geographical region as stated in the Text. See Extended Data Fig. 2 for weekly percent increase in mortality. See Extended Data Fig. 5 for results for US states. In some countries, there was a reduction in mortality relative to a no-pandemic baseline in some weeks, shown as negative numbers. The country’s total excess death toll is the net effect of these reductions and increases in other periods, with all bars adding to 100%.

The subsequent wave(s) of the pandemic (mid-September 2020 to mid-February 2021) saw yet more changes in excess deaths patterns across countries. While New Zealand, Australia, Iceland, Finland, Norway and South Korea remained resilient to the rise in mortality (i.e., no or <2% increase in mortality compared to the no-pandemic baseline), many countries in Europe, especially in Central Europe, experienced a rise in mortality compared to the no-pandemic baseline: by >40% in Slovakia, Czechia and Poland, and by 20-40% in England and Wales, Italy, Austria, Hungary, Montenegro, Croatia, Portugal, Switzerland, Romania, Lithuania, Bulgaria and Slovenia, all with posterior probabilities of positive excess mortality greater than 99%. Excess deaths also reappeared in other countries that had experienced a medium to large toll in the first wave including Belgium, Spain, Scotland, Northern Ireland, Sweden, Canada, France and the Netherlands – some at the same level (France and Northern Ireland) and others at lower levels (Canada, Scotland, Spain, Belgium, Sweden) than the first wave but all lasting for many weeks during this period. The USA had an even larger increase in mortality compared to the no-pandemic baseline after mid-September than it had in the first wave and summer months, making it the only country to maintain a steady burden of excess mortality. There were nonetheless variations in excess deaths over time across different states in the USA (Extended Data Fig. 5).

As a result of these heterogeneous dynamics, there was virtually no correlation between excess mortality in the first wave and the summer period among countries (correlation coefficient of percent increase in the two periods = 0.04), and weakly negative correlation between excess mortality in the first wave and mid-September and later (correlation coefficient = -0.13). This was translated to a variable distribution of excess mortality burden across the three periods (Fig. 3B). For example, the first wave accounted for over half of excess deaths in Scotland, Spain, England and Wales, Canada, Sweden, Belgium and Netherlands. At the other extreme, the period between mid-September 2020 and mid-February 2021 accounted for over 90% of excess deaths in Bulgaria, Croatia, Czechia, Hungary, Latvia, Montenegro, Poland, Slovakia and Slovenia. A similar variation was seen across the US states, with excess deaths along the north-eastern coast (Massachusetts, New Jersey, Connecticut, New York and District of Columbia) being dominated by the first wave, in some southern states (Florida, Arizona, Texas and South Carolina) by the summer, and in the northern plains (Wisconsin, North and South Dakota and Montana) by the post-September period.

Countries differed in how excess deaths were distributed across age groups (Fig. 4). In Denmark, Sweden, France, Switzerland, Belgium and Slovenia >95% of all excess deaths were in those aged 65 years and older. On the other hand, Estonia, Finland (which had the smallest detectable excess mortality of any country), Canada, Lithuania and Chile had the largest share of excess deaths in people aged younger than 65 years. Of the 35 countries with a detectable increase in mortality (defined as median estimated increase of >2%) and sufficient data to analyse by age group, Canada experienced the largest share of excess deaths in those aged younger than 45 years (14% of all excess deaths, followed by the USA and Finland (noting that excess death rates in Finland, although detectable, were lower than in other countries). The high mortality toll in younger Canadians may have been due to Covid-19 death at home^10^ and an increase in deaths from drug overdose ^11^. This division arises largely from how much specific segments of the society, such as workers or care home residents, were exposed to infection. Percent increase in mortality was similar between men and women in most countries (Extended Data Fig. 6). There were nonetheless some exceptions, e.g. in Montenegro, Serbia and the Netherlands deaths increased by a larger percent in men (12%-13%) than women (6%-8%); in contrast, in Slovenia, women (15%) experienced a slightly larger percent increase than men (14%).

**Fig. 4.**
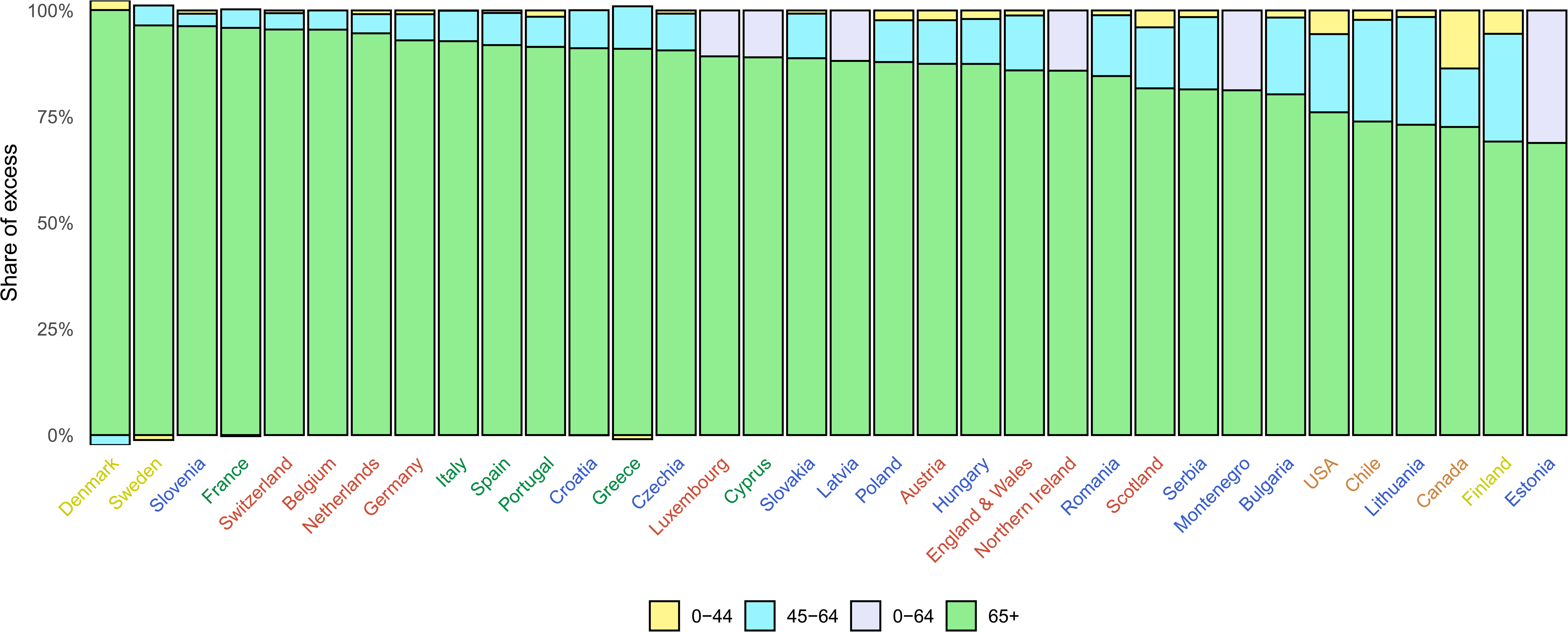
Distribution of excess deaths due to the Covid-19 pandemic by age group. The figure shows the share of excess deaths in each age group by country. There are no bars for Australia, New Zealand, Norway, Iceland and South Korea because we estimated no detectable excess deaths or a potential reduction in mortality compared to the no-pandemic baseline. There is no bar for Malta because we only made all-age estimates for reasons described in Methods. For Luxembourg, Cyprus, Latvia, Northern Ireland, Montenegro and Estonia, analysis was done for 0-64 years without a further split into 0-44 years and 45-64 years for reasons described in Methods.

## Implications for 2021-2022

The magnitude of excess mortality in the first wave of the Covid-19 pandemic was related to two factors. First, how well countries, and subnational entities such as US states, managed the early months of the pandemic – specifically the agility of imposing timely lockdown measures and border controls (e.g., complete or partial travel restrictions and/or quarantine for travellers) and adequate and effective testing, contact tracing and isolation of infected individuals and their contacts, and second, how prepared and resilient the health and social care system was to control the spread of infection, in the community as well as in health facilities and care homes, while continuing routine care^1, 12–17^.

Countries eased or maintained travel restrictions and distancing measures of the first wave to different extents and at different paces^5, 18^. They also differed in terms of testing for surveillance and identifying infected individuals, how well and how fast they traced contacts, and how they supported the isolation of infected individuals and their contacts. Australia and New Zealand took advantage of being islands and pursued an approach of disease elimination^19^ – following strict lockdowns they imposed tight border control which kept cases to sporadic small numbers and allowed careful contact tracing and isolation. Iceland, Norway and South Korea did not close their borders but put in place various forms and durations of quarantine/isolation and testing for travellers. They also effectively integrated their well-coordinated public health capabilities^20^ with modern biomedical (e.g., genomics) and digital technologies (e.g., data from credit card transactions, mobile phones and CCTV footage), and did widespread symptomatic and asymptomatic testing to identify, track and isolate infected individuals and their contacts, and to successfully suppress the epidemic^14, 21–26^, with additional restrictions only when there was a surge in infections. All three countries also have a strong healthcare system that continued to provide routine care alongside care for Covid-19 patients.

At the other extreme, many countries in Central and Eastern Europe, which had put strict measures in place and had experienced no detectable excess mortality during the first half of 2020, removed restrictions on travel and social contact in summer of 2020, at times to a greater extent or at a faster pace than their Western European counterparts^18, 27, 28^. With virtually the entire population still susceptible to infection, this set into motion community transmission, which coincided with the introduction of more transmittable variants of SARS-CoV-2 which were not controlled as fast and as strictly as earlier in 2020, leading to their true ‘first wave’ in Autumn 2020 which was equivalent to or worse than those in their Western European counterparts in magnitude and duration (Extended Data Fig. 1 and 2). Some Mediterranean countries, such as Malta and Greece, and Northwestern European countries, such as Austria and Germany, were also largely spared during the first half of 2020, only to see an increase in deaths in autumn and winter, due to a combination of (tourism-related) travel and increased local mobility and social interactions^29^.

Between these extremes, other countries in Europe and Canada increased their testing capacity, mandated or encouraged masks and face coverings, continued some forms of distancing measures (including occasional lockdowns) and restarted some routine healthcare. There were also improvements in treatments and protocols following large-scale trials and analyses of routine care data^30–32^. These changes meant that, despite the repeated rise in infections, the mortality toll from Covid-19 and other diseases was lower than the first wave but nonetheless considerable in these countries^30^. The continued death toll in these countries may have been because distancing measures were not as stringent as those in the first wave, and because testing, contact tracing and isolation support did not reach the coverage or depth needed to contain transmission, as did those in Iceland and South Korea^22, 33^. This was compounded by more transmittable variants and that the second wave occurred in winter when more time is spent indoors with less ventilation. The experience of the USA did not resemble that of any of the other countries. Rather, different states saw a rise in infections and deaths at different times^34^, because there was little coordinated national response and because periods of extensive travel, such as Thanksgiving and Christmas holidays, led to spread of infection across states.

The observed patterns of excess mortality in the first year of the pandemic indicates that the pandemic’s death toll in the next year is likely to depend on three factors: The first, and most important factor in the countries analysed here will be the breadth and pace of vaccination, including whether vaccination is extended to school-aged children and the use of boosters to enhance immunity especially against new variants of SARS-CoV-2, because vaccines have been shown to be highly effective in preventing (severe) Covid-19 and deaths in trials and in real-world settings^35–37^. Even with high vaccine coverage, some adherence to other measures may be needed when the number of infections rises, because vaccine efficacy is less than 100% and because the morbidity and longer-term health morbidity impacts of infection may be significant. Second, as the direct impacts of the Covid-19 pandemic are reduced through vaccination, the indirect impacts will become more visible. These include how much the backlog of routine care and persistently high health system pressure impacts deaths from other conditions, and the impacts on jobs and income. Mitigating these requires economic and social policies that generate secure employment and income support, and strengthening health and social care. A third, and perhaps more uncertain factor, is the magnitude of direct Covid-19 deaths that might be expected in (northern hemisphere) winter 2021-2022 because retraction of non-pharmaceutical interventions before the entire population is vaccinated may lead to circulating SARS-CoV-2 infections in countries as a whole as well as in specific geographical and sociodemographic subgroups of the population. In mid-February 2021, vaccination rates were still low in the countries included in our analysis, with the highest rates in the UK (22% of adults with one dose and 1% with two doses), Serbia (12% and 3%, respectively), the USA (11% and 4%, respectively) and Chile (11% and 0.3%, respectively). Since then, vaccination accelerated in industrialised countries and emerging economies but is not yet at the levels needed for population immunity to interrupt community transmission. Further, for much of the world, especially in many low and middle-income countries, where access limits the pace of vaccination, the remainder of 2021 and 2022 could look as it did for the countries in this paper over the past year: a combination of extended lockdowns and a large death toll. To avoid this, vaccine roll out must be accompanied with effective actions to both delay and contain infections, especially new variants of concern – through a combination of travel restrictions and isolation of travellers, and effective testing, contact tracing and isolation support.

## Methods

### Data sources

We included industrialised countries in our analysis if:

- Their total population in 2020 was more than 100,000. We excluded countries (e.g., Liechtenstein) with data but with smaller populations because, in many weeks, the number of deaths would be small or zero. This would, in turn, lead to either large uncertainty that would make it hard to differentiate between those places with and without an effect or unstable estimates because the model is fitted to many weeks with zero deaths.
- We could access up-to-date weekly data on all-cause mortality divided by age group and sex that extended through February 2021.
- The time series of data went back at least to the beginning of 2016 so that model parameters could be reliably estimated. For countries with longer time series, we used data starting in 2010.

The sources of population and mortality data are provided in Extended Data Table 2. We calculated weekly population through interpolation of yearly population, consistent with the approach taken by national statistical offices for intra-annual population calculation^38^. Population for 2020 and 2021, where not available, was obtained through linear extrapolation from the last five years. We obtained data on temperature from ERA5^39^, which uses data from global in situ and satellite measurements to generate a worldwide meteorological dataset, with full space and time coverage over our analysis period. We used gridded temperature estimates measured four times daily at a resolution of 30 km to generate weekly temperatures for each first-level administrative region, and gridded population data (https://sedac.ciesin.columbia.edu/data/collection/gpw-v4) to generate population estimates by first-level administrative region in each country. We weighted weekly temperature by population of each first-level administrative region to create national level weekly temperature summaries.

### Statistical methods

The total mortality impact of the Covid-19 pandemic is the difference between the observed number of deaths from all causes of death and the number of deaths had the pandemic not occurred, which is not directly measurable. The most common approach to calculating the number of deaths had the pandemic not occurred has been to use the average number of deaths over previous years, e.g., the most recent five years, for the corresponding week or month when the comparison is made. This approach however does not take into account long- and short-term trends in mortality or time-varying factors like temperature, that are largely external to the pandemic, but also affect death rates.

We developed an ensemble of 16 Bayesian mortality projection models that each make an estimate of weekly death rates that would have been expected if the Covid-19 pandemic had not occurred. We used multiple models because there is inherent uncertainty in the choice of model that best predicts death rates in the absence of pandemic. These models were formulated to incorporate features of weekly death rates, and how they behave in the short-term (week to week) and medium-term (year to year), as follows:

- First, death rates may have a medium-to-long-term trend^40^ that would lead to a lower or higher mortality in 2020-2021 compared to earlier years. Therefore all models included a linear trend term over weekly death rates.
- Second, death rates have a seasonal pattern^41–44^. We included weekly random intercepts for each week of the year. To account for the fact that seasonal patterns “repeat” (i.e., late December and early January are seasonally similar) we used a seasonal structure^45, 46^ for the random intercepts. The seasonal structure allows the magnitude of the random intercepts to vary over time, and implicitly incorporates time-varying factors such as annual fluctuations in flu season.
- Third, death rates in each week may be related to rates in preceding week(s), due to short-term phenomena such as severity of the flu season. We formulated four sets of models to account for this relationship. The weekly random intercepts in these models had a first, second, fourth or eighth order autoregressive structure^45, 46^ The higher-order autoregressive models allow death rates in any week to be informed by those in a progressively larger number of preceding weeks. Further, trends not picked up by the linear or seasonal terms would be captured by these autoregressive terms.
- Fourth and additionally, mortality in one year may depend on mortality in the previous year, in a different way for each month, because phenomena such as seasonal flu may lead to longer-term dependencies in mortality. To allow for this possibility, we used two sets of models, with and without a (first order) autoregressive term over years for each month.
- Fifth, beyond having a seasonal pattern, death rates depend on temperature, and specifically on whether temperature is higher or lower than its long-term norm during a particular time of year^47–52^. The effect of temperature on mortality varies throughout the year, and may be in opposite directions for different times of year. We used two sets of models, one without temperature and one with a weekly term for temperature anomaly, defined as deviation of weekly temperature from the local average weekly temperature over the entire analysis period.
- Finally, death rates may be different around major holidays such as Christmas and New Year either because of changes in human activities and behaviour or, for the countries whose data are registration based, because of delays in registration. We included effects (as fixed intercepts) for the weeks containing Christmas and New Year in all countries. For England and Wales, Scotland and Northern Ireland, we also included effects for the week containing and the week after other public holidays, because reported death rates in weeks that contain a holiday were different from other weeks. This term was tested but not included for other countries because the effect was negligible.

These choices led to an ensemble of 16 Bayesian models (2 yearly autoregressive options x 4 weekly autoregressive options x 2 temperature anomaly options). The ensemble of models is shown in Extended Data Table 5. In each model, the number of weekly deaths follows a Poisson distribution:

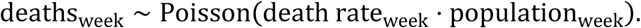

Log-transformed death rates were modelled as a sum of components described above:

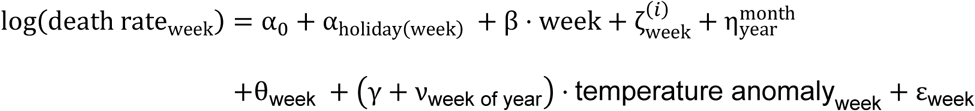

The term α_0_ denotes the overall intercept and α_holiday(week)_ is the holiday intercept, applied to weeks with a holiday. For example, if a week includes the 25^th^ of December then α_holiday(week)_ = α_Christmas_. For weeks that did not contain a holiday, this term did not appear in the above expression. All intercepts were assigned 𝒩(0,1000) priors. The termβ was represents the linear time trend. The coefficient β was also assigned a 𝒩(0,1000) prior.

The models used different orders (first, second, fourth or eighth) of the autoregressive term 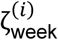 with the superscript *i* denoting the order for weekly mortality patterns. The first-order autoregressive term is defined as 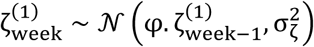 where the parameter φ lies between -1 and 1 and captures the degree of association between the number of deaths in each week and the preceding week. Hyperpriors are placed on the parameters 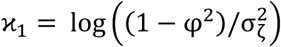 and 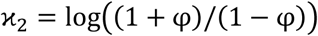 which were assigned logGamma(0.001,0.001) and 𝒩(0,1) distributions respectively. Similarly, an *i^th^* order autoregressive term is given 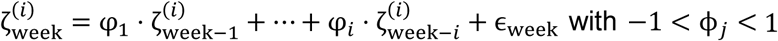 1. The parametrisation of these models was based on the partial autocorrelation function of the sequence ϕ_*j*_^53^.

The term 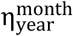 is an autoregressive term of order 1 over years and independent across months, indexed to the month and year to which each particular week belongs. For each month, the autoregressive prior for 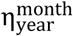 was the same as that for 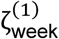 described above. As described above, this term appeared in half of our models.

The term *θ*_week_ captures seasonality in mortality trends with a period of 52 weeks. The sums of every 52 consecutive terms *θ*_week_ + *θ*_week1_ + … *θ*_week+51_ were modelled as independent Gaussian with zero mean and variance 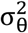. We used a logGamma(0.001, 0.001) prior on the log precision 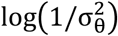. Each week is assigned an index between 1 and 52 depending on which week of the current year it is (the incomplete week 53 is mapped to either index 1 or 52 depending on whether it has greater overlap with week 52 of the current year or week 1 of the following year).

The effect of temperature anomaly on death rates is captured by the two terms γ and ν_week of year_ . The term γ ⋅ temperature anomaly_week_ is the overall association between (log-transformed) death rates and temperature anomaly in a week. The term ν_week of year_ ⋅ temperature anomaly_week_ captures deviations from the overall association for each week of the year. It consists of 52 terms with an independent and identically distributed prior defined via 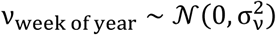, and log-precision 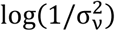 ∼ logGamma(0.001,0.001).

Finally, the term ε_week_ is a zero-mean term that accounts for additional variability. It is assigned an independent and identically distributed prior 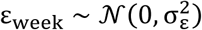, and a logGamma(0.001, 0.001) prior was placed on the log precision 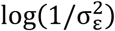. The components 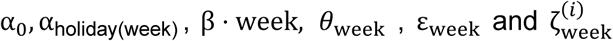 (for autoregressive order *i* = 1, 2, 4 or 8) appear in the expression for log(death rate_week_) in all models. The remaining components appear in some models only. Extended Data Table 5 shows the terms included in each of the 16 models in the ensemble.

We used data on weekly deaths from the start of time series through mid-February 2020 to estimate the parameters of each model, which were then used to predict death rates for the subsequent 52 weeks as estimates of the counterfactual death rates if the pandemic had not occurred. For the projection period, we used recorded temperature so that our projections take into consideration actual temperature in 2020-2021. This choice of training and prediction periods assumes that the number of deaths that are directly or indirectly related to the Covid-19 pandemic was negligible through mid-February 2020 in these countries^1^, and separates the training data from subsequent weeks when impacts may have appeared.

All models were fitted using integrated nested Laplace approximation (INLA)^54^, implemented in the R-INLA software (version 20.03). We used a model averaging approach to combine the predictions from the 16 models in the ensemble^55, 56^. Specifically, we took 2,000 draws from the posterior distribution of age-specific deaths under each of the 16 models, and pooled the 32,000 draws to obtain the posterior distribution of deaths if the Covid-19 pandemic had not taken place. This approach generates a distribution of estimates that has equal samples from each model in the ensemble, and hence incorporates both the uncertainty of estimates from each model and the uncertainty in the choice of model. The reported credible intervals represent the 2.5^th^ and 97.5^th^ percentiles of the resultant posterior distribution of the draws from the entire ensemble. We also report the posterior probability that an estimated increase (or decrease) in deaths corresponds to a true increase (or decrease). Posterior probability represents the inherent uncertainty in how many deaths would have occurred in the absence of the pandemic. In a country and week in which the actual number of deaths is the same as the posterior median of the number expected in a no-pandemic counterfactual, an increase in deaths is statistically indistinguishable from a decrease; in such a situation, there is a 50% posterior probability of an increase and a 50% posterior probability of a decrease. Where the entire posterior distribution of the number of deaths expected without the pandemic is smaller than the actual number of deaths, there is a ∼100% posterior probability of an increase and a ∼0% posterior probability of a decrease and vice versa. For most countries and weeks, the posterior distribution of the number of deaths expected without the pandemic covers the observed number, but there is asymmetry in terms of whether much of the distribution is smaller or larger than the observed number. In such cases, there would be uneven posterior probabilities of an increase versus decrease in deaths, with the two summing to 100% (for example, 80% and 20%). Posterior probabilities more distant from 50%, toward either 0% or 100%, indicate more certainty.

We did all analyses separately by sex and age group (0-44 years, 45-64 years, 65+ years) for countries with 2020 population of at least two million, where age- and sex-specific data were available (Extended Data Table 2). For countries with 2020 population less than 2 million, we did our analyses for two age groups (0-64 years and 65+ years) because, in many weeks, the number of deaths in the age group 0-44 would be small or zero, which would lead to either large uncertainty or unstable estimates. For the same reason, for countries with population under 500,000 (Iceland and Malta), we did our analyses for both sexes and all age groups combined. Models were also run for all ages and both sexes combined; the posterior median of resultant estimates were nearly identical to the sum of the age-sex-specific ones, with a mean relative difference of 0.2%, ranging from -1.7% to 1.1%. For this reason, in figures and tables that are for all ages and both sexes, we report results from the combined model so that the uncertainty of the estimates is correctly reported.

### Validation of no-pandemic counterfactual weekly deaths

We tested how well our model ensemble estimates the number of deaths expected had the pandemic not occurred by withholding data for 52 weeks starting from mid-February (i.e., the same projection period as done for 2020-2021) for an earlier year and using the preceding time series of data to train the models. In other words, we created a situation akin to 2020-2021 for an earlier year. We then projected death rates for the weeks with withheld data, and evaluated how well the model ensemble projections reproduced the known-but-withheld death rates. We repeated this for three different periods: 2017-2018 (i.e., train model using data from January 2010 to mid-February 2017 and test for the subsequent 52 weeks), 2018-2019 (i.e., train model using data from January 2010 to mid-February 2018 and test for the subsequent 52 weeks), and 2019-2020 (i.e., train model using data from January 2010 to mid-February 2019 and test for the subsequent 52 weeks). We performed these tests for each country using data for both sexes and all ages. We report the projection error (which measures systematic bias) and absolute projection error (which measures any deviation from the data). Additionally, we report coverage of the projection uncertainty; if projected death rates and their uncertainties are well estimated, the estimated 95% credible intervals should cover 95% of the withheld data.

The results of model validation (Extended Data Table 3) show that the estimates of how many deaths would be expected had the pandemic not occurred from the Bayesian model ensemble were unbiased, with mean relative projection errors of 1.5% (between 0.6% and 2.2% in different years). The mean relative absolute error was between 8.0% and 8.7% in different years. 95% coverage, which measures how well the posterior distributions of projected deaths coincide with withheld data was 96% for all years, which shows that the posterior distribution is well estimated.

### Strengths and limitations

The main strength of our work is the development and application of a method to systematically and consistently use time series data from previous years to estimate how many deaths would be expected in the absence of pandemic through early 2021. The models incorporated important features of mortality, including seasonality of death rates, how mortality in one week or year may depend on previous week(s) and year(s), and the seasonally-variable role of temperature. To our knowledge, our models are the only ones that formally incorporated the role of temperature on weekly mortality, and accounted for dependency of mortality in one week on preceding week(s) and in one year on preceding year(s). This methodology allows more robust estimation of the total impacts of the pandemic, especially as more time elapses since the beginning of the pandemic. It also enables comparisons of excess deaths across countries on a real-time basis. By modelling death rates, rather than simply the number of deaths as is done in most other analyses, we account for changes in population size and age structure. We used an ensemble of models which typically leads to more robust projections and better accounts for both the uncertainty associated with each individual model and that of model choice. As a result, our approach gives a more complete picture of the inherent uncertainty in how many excess deaths the pandemic has caused than approaches that are not probabilistic or use a single model.

A limitation of our work is that we did not have data on underlying cause of death. Having a breakdown of deaths by underlying cause will help develop cause-specific models and understand which causes have exceeded or fallen below the levels expected. Nor did we have data on total mortality by individual or community sociodemographic status to understand inequalities in the impacts of the pandemic beyond deaths assigned to Covid-19 as the underlying cause of death. Where data have been analysed for population subgroups, excess mortality tends to be higher in marginalised individuals and communities^57–59^. More detailed data will allow more granular analysis of the impacts of the pandemic, which can in turn inform resource allocation and a more targeted approach to mitigating both the direct and indirect effects of the Covid-19 pandemic.

### Comparison with other estimates

*Financial Times* and *The Economist*’s excess deaths tracker report the number of excess deaths for various countries based on comparisons of deaths in 2020 and 2021 with 2015-2019 averages. This approach does not account for general trends in mortality nor for factors like temperature that affect mortality and vary from year to year. *The Economist* has also recently published a set of excess deaths estimates using data from the Human Mortality Database and the World Mortality Dataset, and an ensemble of gradient boosted decision trees. Countries with small, medium and large number of excess deaths are largely consistent between our analysis and these sources. There are nonetheless some differences. For example, we estimated ∼621,000 excess deaths for the US, compared to ∼549,000 by *Financial Times* and ∼600,000 by *The Economist* (for comparison, US CDC estimated 646,000 excess deaths). Our median excess death estimate for Denmark was about twice as large as that of *Financial Times*, and those for Greece and Serbia about one third smaller. Similarly, *The Economist* model predicted a mortality deficit of about 3,900 deaths for South Korea and a small but positive number of excess deaths in Australia (∼1,600 deaths), while our estimate is that there was no detectable change in mortality in South Korea and a deficit of over 4,500 deaths in Australia. Nonetheless, the 95% credible interval of our estimates contained those of *Financial Times* and *The Economist*.

The Institute for Health Metrics and Evaluation has released numbers of “total Covid-19 deaths” by fitting a model for seasonality (the details of seasonal model are not currently available) and projecting the residuals for pre-2020 using a spline model. The models do not account for temperature, as ours do, but hot summer weeks with particularly large deaths were excluded. Several sources have commented that the estimates are likely an overestimate ^60–63^. For example the Institute estimated ∼138,000 deaths for the UK and ∼760,000 for the USA for the same period as our analysis, compared to ∼111,000 and ∼621,000 by us (for comparison, UK national statistical offices estimated ∼118,500 for England, Wales, Scotland and Northern Ireland; US CDC estimated ∼646,000). They estimated ∼35,000 deaths for Canada, compared to ∼15,000 by us and ∼19,000 by Statistics Canada, and ∼38,000 excess deaths for Portugal, compared to ∼21,000 by us. EuroMoMo fits a sinusoidal seasonal model to death counts but does not report country-specific excess deaths and hence could not be compared with our results.

The UK Office for National Statistics (ONS) calculated a number of age-standardised measures of excess mortality for 15 European countries based on comparisons of deaths in 2020 with 2015-2019 averages ^64^, as did Eurostat for the monthly number of deaths. These analyses did not account for temperature and holidays, and the Eurostat analysis did not account for changes in population. The ONS concluded that Norway, Finland, Denmark and Latvia, Cyprus and Estonia had a mortality deficit whereas our estimates indicated no detectable excess mortality for Norway, and increases from 2 to 8% for the other countries. Differences between our results and those of the ONS may be partly related to the fact that ONS analysis also included the pre-pandemic months and did not account for interannual variations in temperature. For example, in the northern hemisphere, the first and last three months of 2020 were on average warmer than the average of the past five years but weeks 13-40 were on average slightly cooler.

## Data availability

Estimates of weekly excess deaths by country will be available from http://globalenvhealth.org/code-data-download/ upon publication of the paper. Input data on deaths, population and temperature will also be available from http://globalenvhealth.org/code-data-download/.

## Code availability

The computer code for the Bayesian model ensemble used in this work will be available at http://globalenvhealth.org/code-data-download/ upon publication of the paper.

## Author contributions

ME, VK and JEB designed the study. VK and JEB developed and tested statistical methods with input from TR, RMP, MG and ME. VK wrote computer code, conducted analysis and prepared results. VK, RMP, JEB and Y-HK accessed, harmonised and analysed data. ME and VK wrote the first draft of the paper and other authors contributed to the paper.

## Author information

ME reports a charitable grant from the AstraZeneca Young Health Programme, outside the submitted work. JP-S is vice-chair of the Royal Society for Public Health and reports personal fees from Novo Nordisk A/S and Lane, Clark & Peacock LLP, outside of the submitted work.

## Acknowledgements

The development of methodology for estimating the impact of pandemic as an extreme event was supported by the Pathways to Equitable Healthy Cities grant from the Wellcome Trust [209376/Z/17/Z]. Work on the US mortality data was partially supported by a grant from the US Environmental Protection Agency (EPA), as part of the Center for Clean Air Climate Solution (assistance agreement no. R835873). This article has not been formally reviewed by the EPA. The views expressed in this document are solely those of the authors and do not necessarily reflect those of the EPA. The EPA does not endorse any products or commercial services mentioned in this publication. Work on the UK mortality data was partially supported by the British Heart Foundation (Centre of Research Excellence grant RE/18/4/34215). For the purpose of Open Access, the author has applied a CC BY public copyright licence to any Author Accepted Manuscript version arising from this submission.

---

**Extended Data Fig. 1.**
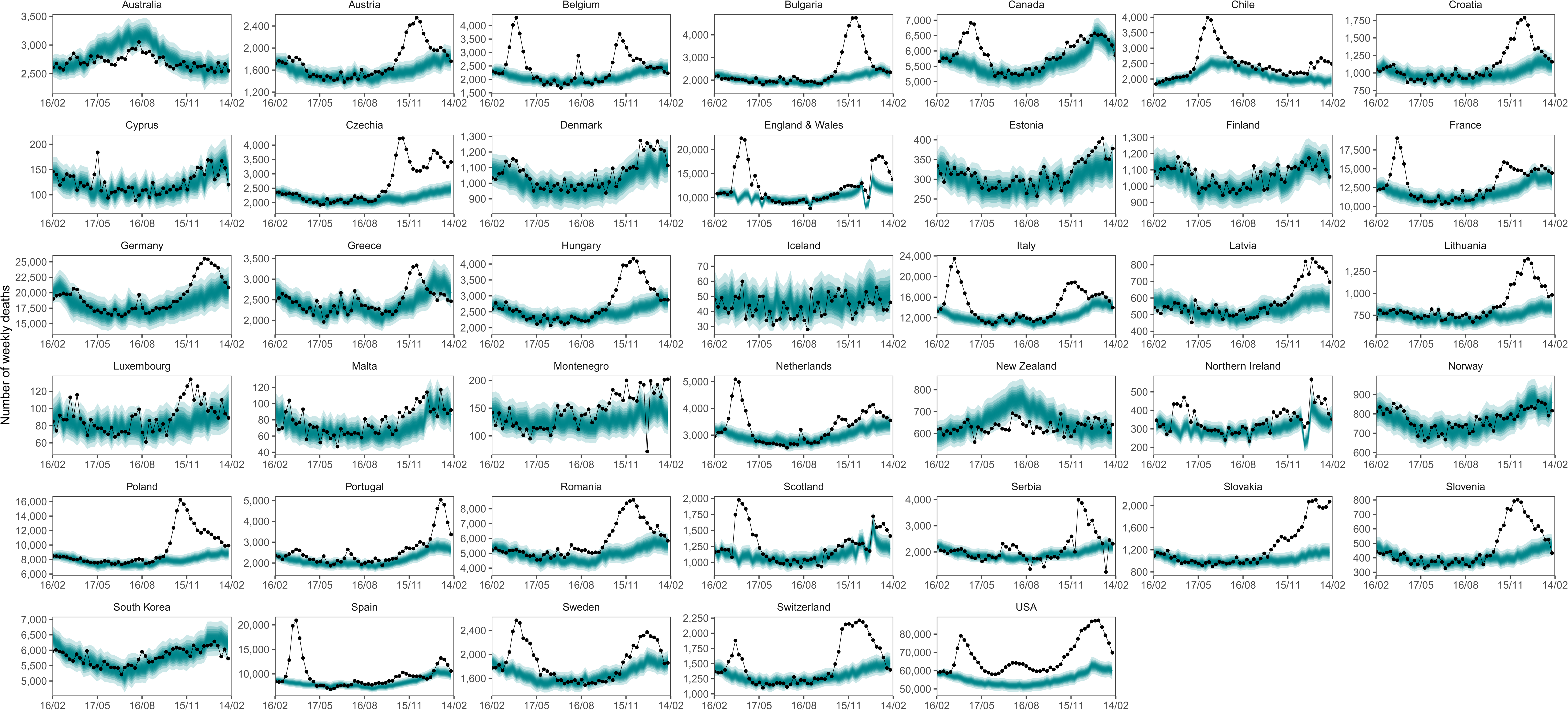
Weekly number of deaths from mid-February 2020 through mid-February 2021. The points show reported deaths. The turquoise shading shows the credible intervals around the median prediction, from 5% (dark) to 95% (light) in 10% increments.

**Extended Data Fig. 2.**
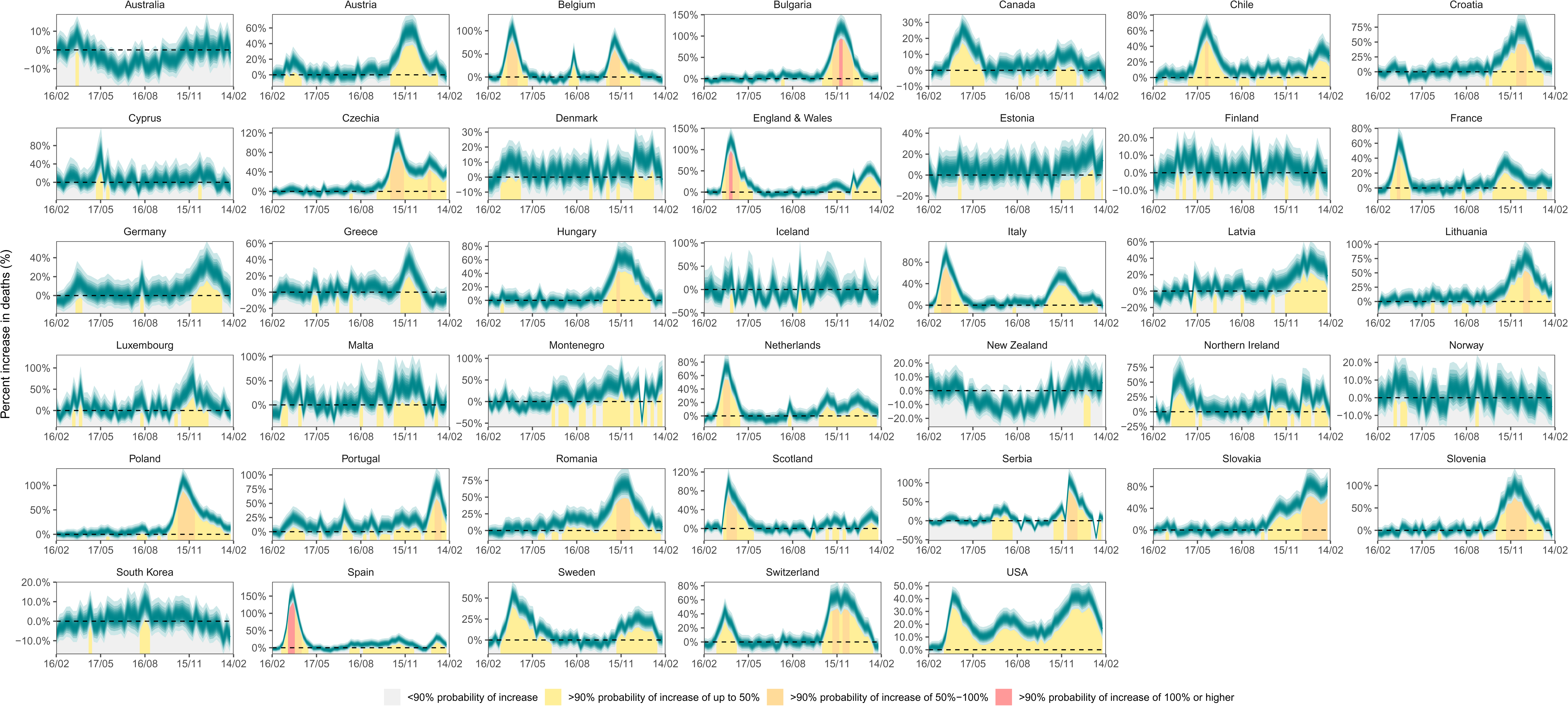
Weekly percent increase in mortality as a result of the Covid-19 pandemic by country. The turquoise shading shows the credible intervals around the median prediction, from 5% (dark) to 95% (light) in 10% increments. The background shading indicates the magnitude of the weekly increase that was detectable with a posterior probability of at least 90%.

**Extended Data Fig. 3.**
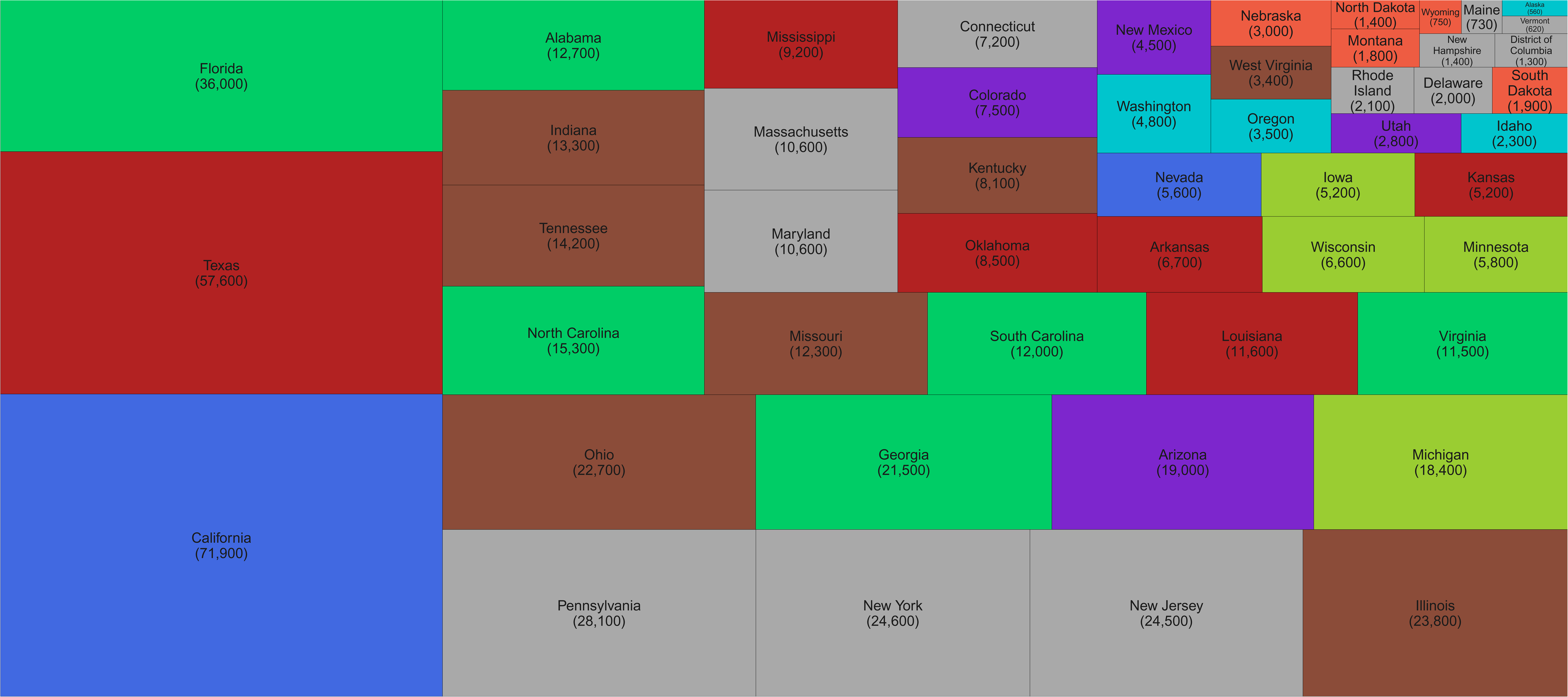
Number of excess deaths due to the first year of the Covid-19 pandemic by US state. The size of each rectangle shows the number of deaths from all causes in excess of what would be expected if there had been no Covid-19 pandemic from mid-February 2020 through mid-February 2021 for each state and the District of Columbia. There is no segment for Hawaii because we estimated no detectable excess deaths. Results for North Carolina are calculated using 50 weeks of data because deaths for February 2021 have not been reported so far.

**Extended Data Fig. 4.**
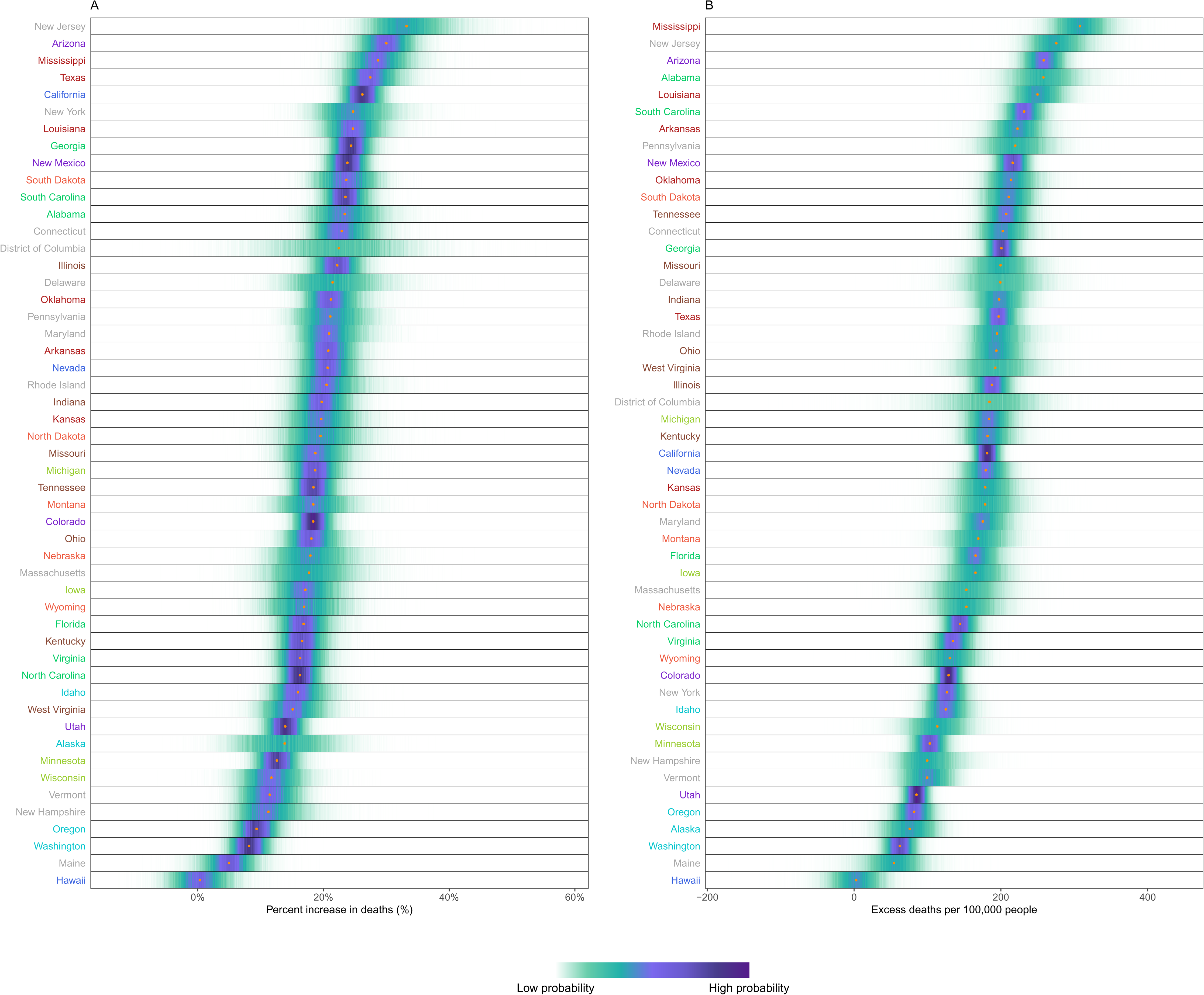
Excess mortality due to the Covid-19 pandemic, by US state. (A) Posterior distribution of percent increase in deaths from any cause from mid-February 2020 to mid-February 2021. Gold dots show the posterior medians. (B) Posterior distribution of excess deaths from any cause per 100,000 people from mid-February 2020 to mid-February 2021. Gold dots show the posterior medians. States are ordered vertically by median increase from smallest (at the bottom) to the largest (at the top).

**Extended Data Fig. 5.**
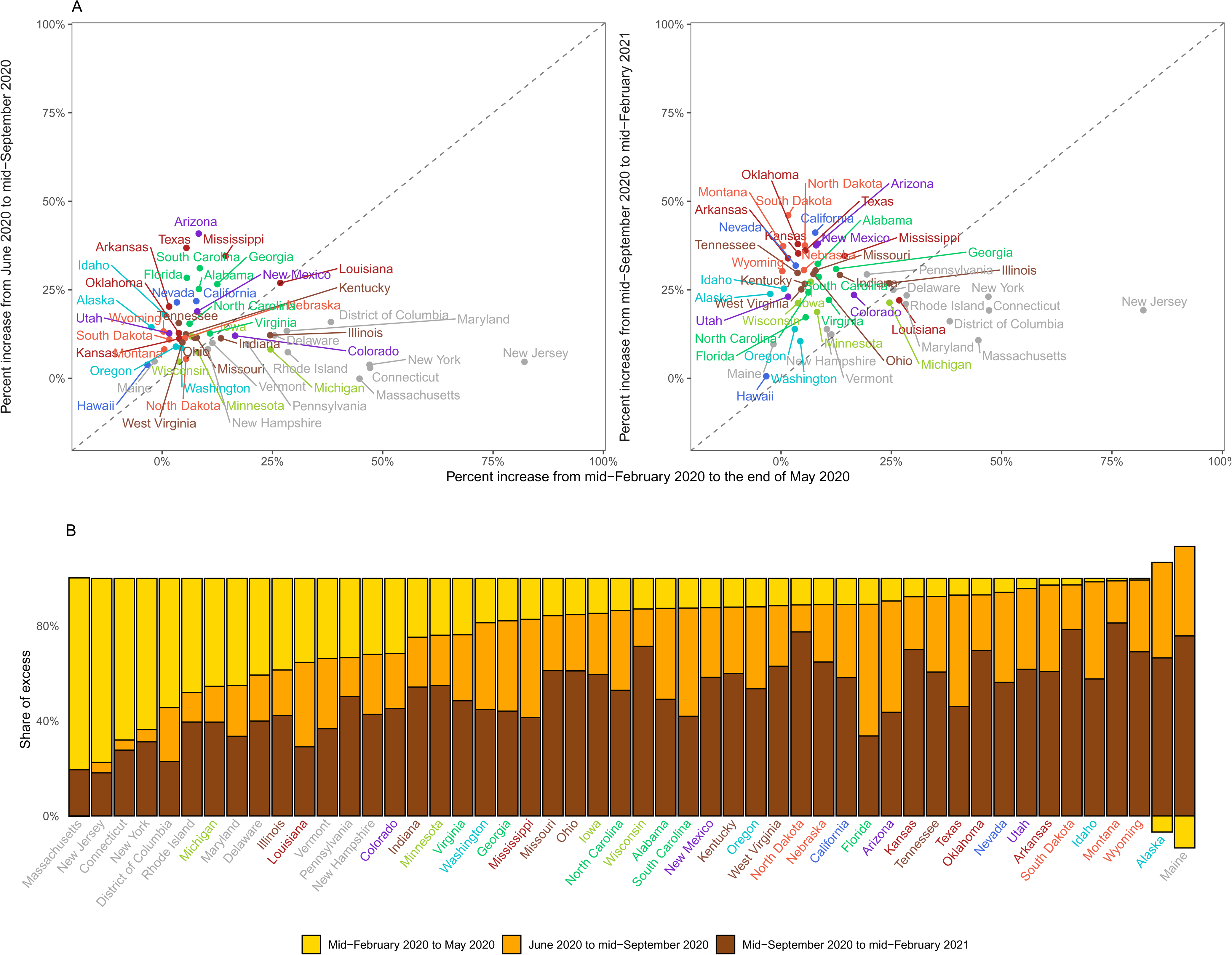
Excess mortality due to the Covid-19 pandemic in different time periods for US States. (A) Comparison of percent increase in mortality from any cause in excess of what would be expected if there had been no Covid-19 pandemic in summer (beginning of June 2020 to mid-September 2020) and subsequent waves (mid-September 2020 to mid-February 2021) with the first wave (mid-February 2020 to end of May 2020) in each state. (B) Proportion of excess deaths in each of the above three periods in each state. There is no bar for Hawaii because we estimated no detectable excess deaths. In some states, there was a reduction in mortality relative to a no-pandemic baseline in some weeks, shown as negative numbers. The state’s total excess death toll is the net effect of these reductions and increases in other periods, with all bars adding to 100%.

**Extended Data Fig. 6.**
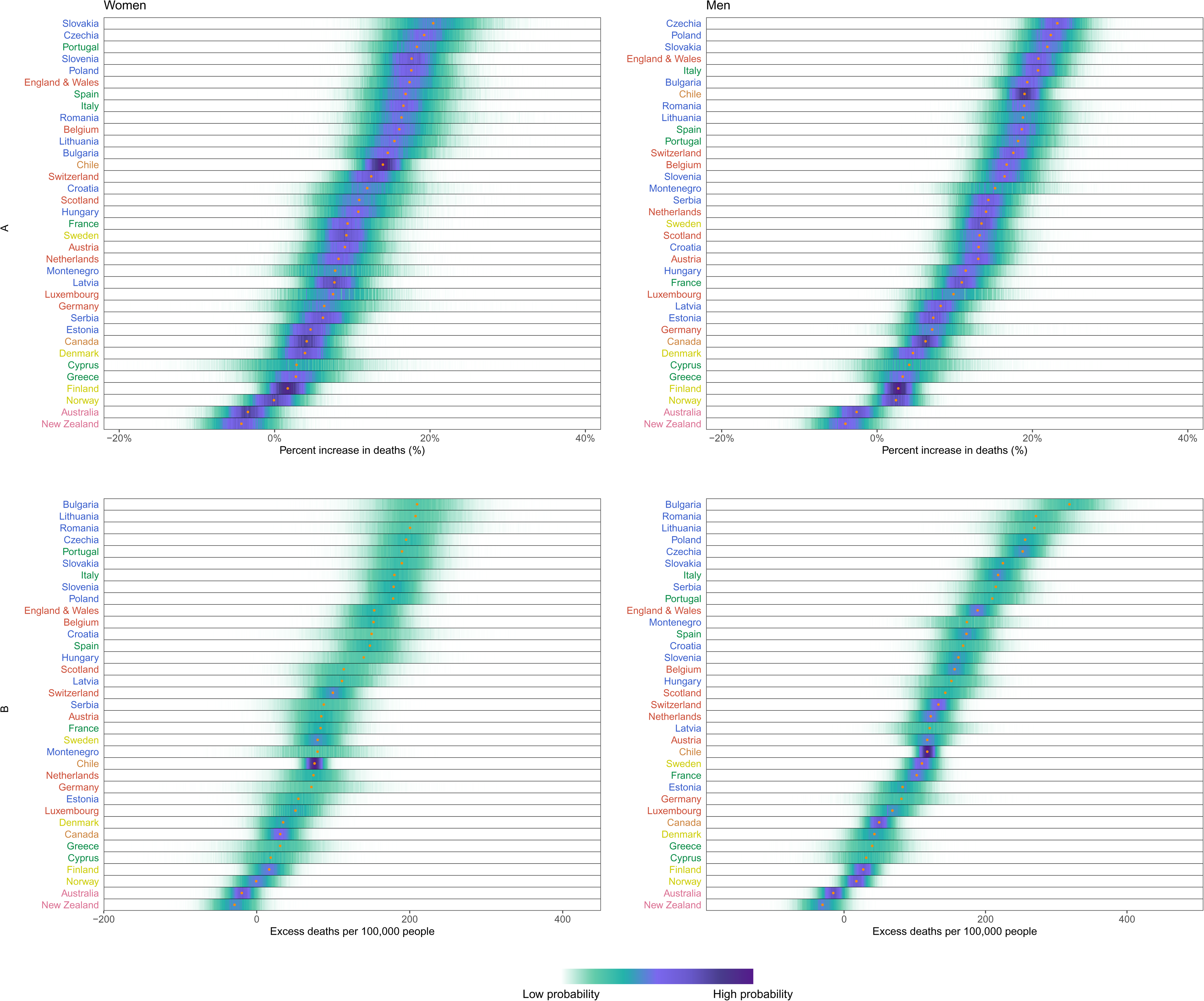
Excess mortality due to the Covid-19 pandemic, by country and sex. (A) Posterior distribution of percent increase in deaths from any cause from mid-February 2020 to mid-February 2021. Gold dots show the posterior medians. (B) Posterior distribution of excess deaths from any cause per 100,000 people from mid-February 2020 to mid-February 2021. Gold dots show the posterior medians. Countries are ordered vertically by median increase from smallest (at the bottom) to the largest (at the top). Data for Northern Ireland, South Korea and USA were only available for both sexes combined and did not allow sex-specific results. There are no segments for Malta and Iceland because estimates for these countries were only made for both sexes combined, for reasons described in Methods.

**Extended Data Table 1.** Number of excess deaths from any cause and deaths assigned to Covid-19 due from mid-February 2020 to mid-February 2021, by country. Excess deaths ≥1,000 are rounded to the nearest hundred and excess deaths <1,000 to the nearest ten. Deaths assigned to Covid-19 were taken directly from the cited sources and not rounded.

| Country | Number of excess deaths (95% credible interval) | Number of deaths assigned to Covid-19 as underlying cause<sup>1</sup> |
| --- | --- | --- |
| Australia | -4,530 (-11,560 to 1,900) | 909 |
| Austria | 9,100 (4,800 to 13,100) | 8,258 |
| Belgium | 17,900 (12,200 to 23,600) | 22,077 |
| Bulgaria | 18,200 (12,700 to 23,300) | 9,854 |
| Canada | 15,400 (4,400 to 27,100) | 21,723 |
| Chile | 18,400 (15,100 to 21,700) | 20,126 |
| Croatia | 6,400 (3,300 to 9,400) | 5,449 |
| Cyprus | 260 (-380 to 870) | 229 |
| Czechia | 23,900 (18,500 to 29,300) | 19,330 |
| Denmark | 2,400 (-150 to 5,200) | 2,343 |
| England and Wales | 102,100 (75,300 to 129,000) | 128,077 |
| Estonia | 920 (270 to 1,600) | 535 |
| Finland | 1,200 (-440 to 2,800) | 756 |
| France | 62,700 (32,600 to 95,300) | 84,306 |
| Germany | 63,900 (-3,940 to 136,200) | 67,903 |
| Greece | 3,700 (-4,250 to 11,100) | 6,297 |
| Hungary | 14,200 (7,300 to 21,200) | 14,347 |
| Iceland | -140 (-340 to 40) | 29 |
| Italy | 118,100 (87,300 to 148,000) | 95,718 |
| Latvia | 2,200 (1,100 to 3,300) | 1,542 |
| Lithuania | 6,600 (4,400 to 8,800) | 3,178 |
| Luxembourg | 390 (140 to 640) | 625 |
| Malta | 320 (20 to 640) | 304 |
| Montenegro | 780 (370 to 1,200) | 950 |
| Netherlands | 17,300 (9,400 to 25,500) | 15,231 |
| New Zealand | -1,370 (-3,300 to 550) | 26 |
| Northern Ireland | 2,300 (1,300 to 3,300) | 2,751 |
| Norway | 480 (-1,090 to 2,100) | 608 |
| Poland | 82,100 (62,700 to 101,100) | 42,171 |
| Portugal | 20,600 (14,100 to 27,000) | 15,962 |
| Romania | 45,400 (30,500 to 61,300) | 19,894 |
| Scotland | 7,100 (3,700 to 10,600) | 9,355 |
| Serbia | 10,300 (5,600 to 14,800) | 4,337 |
| Slovakia | 11,200 (8,100 to 14,100) | 6,577 |
| Slovenia | 3,600 (2,700 to 4,400) | 4,057 |
| South Korea | 390 (-11,150 to 10,800) | 1,562 |
| Spain | 76,000 (52,800 to 100,000) | 67,636 |
| Sweden | 9,900 (6,400 to 13,600) | 12,911 |
| Switzerland | 10,100 (7,000 to 13,200) | 9,174 |
| USA | 621,100 (520,100 to 749,700) | 526,729 |

<sup>1</sup> Data are from Office for National Statistics for England and Wales (https://www.ons.gov.uk/peoplepopulationandcommunity/birthsdeathsandmarriages/deaths/datasets/weeklyprovisionalfiguresondeathsregisteredinenglandandwales), NRS for Scotland (https://www.nrscotland.gov.uk/covid19stats), NISRA for Northern Ireland (https://www.nisra.gov.uk/system/files/statistics/Weekly_Deaths%20-%20w%20e%204th%20June%202021.XLSX) and the European Centre for Disease Prevention and Control (ECDC) for other countries (https://opendata.ecdc.europa.eu/covid19/nationalcasedeath/csv; accessed on 9 July 2021).

**Extended Data Table 2.**
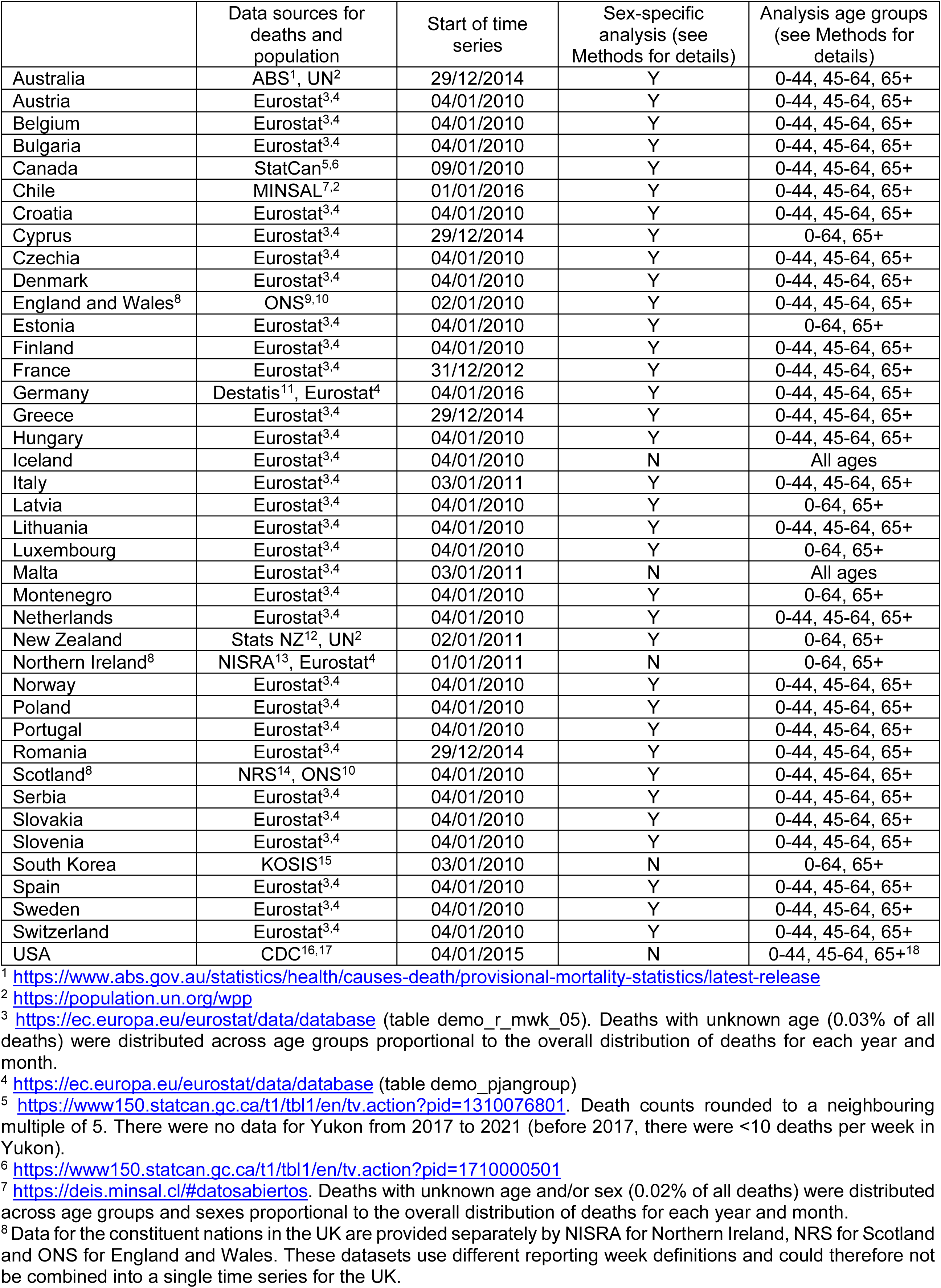
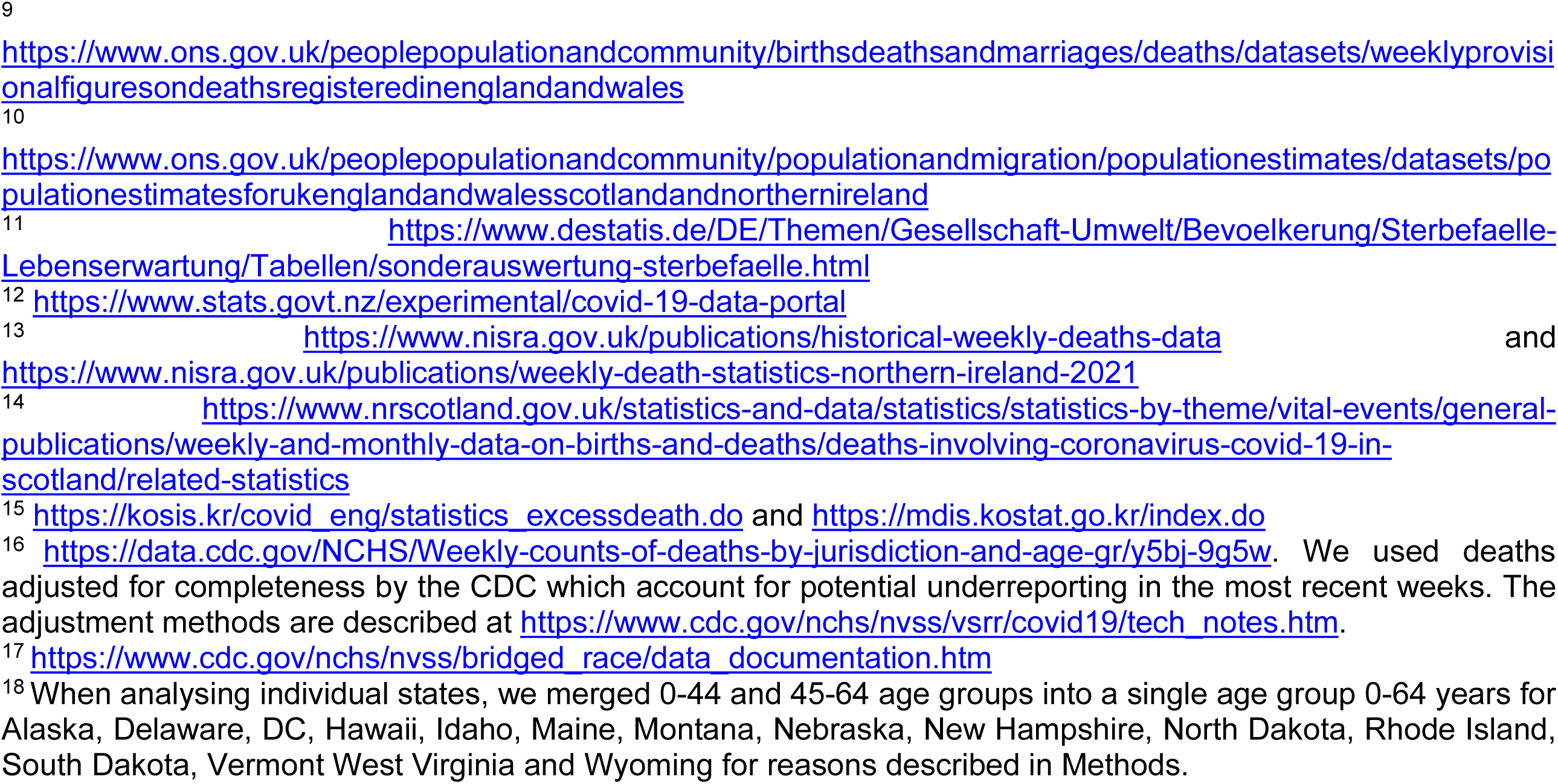
Sources of data on deaths and population.

**Extended Data Table 3.** Results of the external predictive validity (out-of-sample validation) of the estimated no-pandemic counterfactual weekly deaths from the Bayesian model ensemble. Each number represents the total error over the validation period, averaged across countries.

| Validation year | Projection error<br>(relative projection error) | Absolute projection error (relative<br>absolute projection error) | Percent covered by<br>95% credible interval |
| --- | --- | --- | --- |
| 2017 | 1,893 (1.8%) | 9,488 (8.5%) | 97% |
| 2018 | 1,107 (0.5%) | 9,455 (8.7%) | 94% |
| 2019 | 3,306 (2.2%) | 8,645 (8.0%) | 98% |
| All three years | 2,102 (1.5%) | 9,196 (8.4%) | 96% |

\* Australia, Chile, Cyprus, Germany, Greece, Romania and USA were not used for validation analysis because they had shorter time series. Hence leaving out the last three years of data would leave a time series that was too short for estimating model parameters.

**Extended Data Table 4.** Combination of terms used in each of the 16 models for estimating number of weekly deaths that would be expected had the pandemic not occurred. See Methods for an explanation of each term.

| Model number | Global intercepts<sup>1</sup> | Time slope | Non-linear (autoregressive) term over weeks | Seasonal term | Non-linear (autoregressive) month-specific term over years | Temperature anomaly terms |
| --- | --- | --- | --- | --- | --- | --- |
| 1 | $\alpha_0 + \alpha_{\text{holiday(week)}}$ | $\beta \cdot \text{week}$ | $\zeta_{\text{week}}^{(1)}$ | $\theta_{\text{week}}$ | - | - |
| 2 | $\alpha_0 + \alpha_{\text{holiday(week)}}$ | $\beta \cdot \text{week}$ | $\zeta_{\text{week}}^{(1)}$ | $\theta_{\text{week}}$ | - | $(\gamma + v_{\text{week of year}}) \cdot \text{temperature anomaly}_{\text{week}}$ |
| 3 | $\alpha_0 + \alpha_{\text{holiday(week)}}$ | $\beta \cdot \text{week}$ | $\zeta_{\text{week}}^{(1)}$ | $\theta_{\text{week}}$ | $\eta_{\text{year}}^{\text{month}}$ | - |
| 4 | $\alpha_0 + \alpha_{\text{holiday(week)}}$ | $\beta \cdot \text{week}$ | $\zeta_{\text{week}}^{(1)}$ | $\theta_{\text{week}}$ | $\eta_{\text{year}}^{\text{month}}$ | $(\gamma + v_{\text{week of year}}) \cdot \text{temperature anomaly}_{\text{week}}$ |
| 5 | $\alpha_0 + \alpha_{\text{holiday(week)}}$ | $\beta \cdot \text{week}$ | $\zeta_{\text{week}}^{(2)}$ | $\theta_{\text{week}}$ | - | - |
| 6 | $\alpha_0 + \alpha_{\text{holiday(week)}}$ | $\beta \cdot \text{week}$ | $\zeta_{\text{week}}^{(2)}$ | $\theta_{\text{week}}$ | - | $(\gamma + v_{\text{week of year}}) \cdot \text{temperature anomaly}_{\text{week}}$ |
| 7 | $\alpha_0 + \alpha_{\text{holiday(week)}}$ | $\beta \cdot \text{week}$ | $\zeta_{\text{week}}^{(2)}$ | $\theta_{\text{week}}$ | $\eta_{\text{year}}^{\text{month}}$ | - |
| 8 | $\alpha_0 + \alpha_{\text{holiday(week)}}$ | $\beta \cdot \text{week}$ | $\zeta_{\text{week}}^{(2)}$ | $\theta_{\text{week}}$ | $\eta_{\text{year}}^{\text{month}}$ | $(\gamma + v_{\text{week of year}}) \cdot \text{temperature anomaly}_{\text{week}}$ |
| 9 | $\alpha_0 + \alpha_{\text{holiday(week)}}$ | $\beta \cdot \text{week}$ | $\zeta_{\text{week}}^{(4)}$ | $\theta_{\text{week}}$ | - | - |
| 10 | $\alpha_0 + \alpha_{\text{holiday(week)}}$ | $\beta \cdot \text{week}$ | $\zeta_{\text{week}}^{(4)}$ | $\theta_{\text{week}}$ | - | $(\gamma + v_{\text{week of year}}) \cdot \text{temperature anomaly}_{\text{week}}$ |
| 11 | $\alpha_0 + \alpha_{\text{holiday(week)}}$ | $\beta \cdot \text{week}$ | $\zeta_{\text{week}}^{(4)}$ | $\theta_{\text{week}}$ | $\eta_{\text{year}}^{\text{month}}$ | - |
| 12 | $\alpha_0 + \alpha_{\text{holiday(week)}}$ | $\beta \cdot \text{week}$ | $\zeta_{\text{week}}^{(4)}$ | $\theta_{\text{week}}$ | $\eta_{\text{year}}^{\text{month}}$ | $(\gamma + v_{\text{week of year}}) \cdot \text{temperature anomaly}_{\text{week}}$ |
| 13 | $\alpha_0 + \alpha_{\text{holiday(week)}}$ | $\beta \cdot \text{week}$ | $\zeta_{\text{week}}^{(8)}$ | $\theta_{\text{week}}$ | - | - |
| 14 | $\alpha_0 + \alpha_{\text{holiday(week)}}$ | $\beta \cdot \text{week}$ | $\zeta_{\text{week}}^{(8)}$ | $\theta_{\text{week}}$ | - | $(\gamma + v_{\text{week of year}}) \cdot \text{temperature anomaly}_{\text{week}}$ |
| 15 | $\alpha_0 + \alpha_{\text{holiday(week)}}$ | $\beta \cdot \text{week}$ | $\zeta_{\text{week}}^{(8)}$ | $\theta_{\text{week}}$ | $\eta_{\text{year}}^{\text{month}}$ | - |
| 16 | $\alpha_0 + \alpha_{\text{holiday(week)}}$ | $\beta \cdot \text{week}$ | $\zeta_{\text{week}}^{(8)}$ | $\theta_{\text{week}}$ | $\eta_{\text{year}}^{\text{month}}$ | $(\gamma + v_{\text{week of year}}) \cdot \text{temperature anomaly}_{\text{week}}$ |

<sup>1</sup> Due to the short duration of the time series, the holiday term was not identifiable for Chile (in the presence of the seasonal term) and was therefore not included.

